# Antibodies against type-I Interferon: detection and association with severe clinical outcome in COVID-19 patients

**DOI:** 10.1101/2021.04.02.21253262

**Authors:** Goncalves David, Mezidi Mehdi, Bastard Paul, Perret Magali, Saker Kahina, Fabien Nicole, Pescarmona Rémi, Lombard Christine, Walzer Thierry, Casanova Jean-Laurent, Belot Alexandre, Richard Jean-Christophe, Trouillet-Assant Sophie

## Abstract

**Objectives:** Impairment of type I interferon (IFN-I) immunity has been reported in critically-ill COVID-19 patients. This defect can be explained in a subset of patients by the presence of circulating autoantibodies (auto-Abs) against IFN-I. We set out to improve the detection and the quantification of IFN-I auto-Abs in a cohort of critically-ill COVID-19 patients, in order to better evaluate the prevalence of these Abs as the pandemic progresses, and how they correlate with the clinical course of the disease.

**Methods:** The concentration of anti*-*IFN-α_2_ Abs was determined in the serum of 84 critically-ill COVID-19 patients who were admitted to ICU in *Hospices Civils de Lyon*, France using a commercially available kit (Thermo-Fisher, Catalog #BMS217).

**Results:** A total of 21/84 (25%) critically-ill COVID-19 patients had circulating anti-IFN-α_2_ Abs above cut-off (>34 ng.mL^-1^). Among them, 15/21 had Abs with neutralizing activity against IFN-α_2_, *i*.*e*. 15/84 (18%) of critically-ill patients. In addition, we noticed an impairment of the IFN-I response in the majority of patients with neutralizing anti-IFN-α_2_ Abs. There was no significant difference in the clinical characteristics or outcome of with or without neutralizing anti-IFN-α_2_ auto-Abs. We detected anti-IFN-α_2_ auto-Abs in COVID-19 patients’ sera throughout their ICU stay. Finally, we also found auto-Abs against multiple subtypes of IFN-I including IFN-ω.

**Conclusions:** We reported that 18% of critically-ill COVID-19 patients were positive for IFN-I auto-Abs, confirming that the presence of these antibodies is associated with higher risk of developing a criticall COVID-19 form.

## Introduction

Severe acute respiratory syndrome coronavirus-2 (SARS-CoV2) infection leads to coronavirus disease 19 (COVID-19), whose spectrum of clinical presentations is wide and includes severe pneumonia. The anti-SARS-CoV-2 immune response has been extensively studied and defects in antiviral mechanisms have been linked to disease severity. In particular, impairment of type I interferon (IFN-I) immunity has been reported in critically-ill COVID-19 patients. Such defect can be due to either inherited genetic deficiencies in the IFN-I pathway or the occurrence of circulating auto-antibodies (auto-Abs) directed against 14 or the 17 individual IFN-I ^1-4^. These auto-Abs have also been detected in a third of patients from a small international cohort who had suffered from severe adverse events following Yellow Fever vaccination (YFV-17D)^5^. These findings advocate for the development of diagnostic tools for the detection of IFN-I auto-Abs in routine laboratories, in order to identify early patients at risk of developing severe forms of COVID-19 and to analyze the prevalence of IFN-I Abs as the pandemic progresses and the virus evolves. To this aim, we tested a commercially available kit measuring IFN-I auto-Abs levels in the serum of COVID-19 patients.

## Results

A total of 84 critically-ill COVID-19 patients, 11 patients with autoimmune polyendocrinopathy type 1 syndrome (APS-1), 10 mildly-symptomatic COVID-19 healthcare workers, and 76 healthy controls were included in the study. The critically-ill COVID-19 patients were admitted to ICU in the Lyon University Hospital, France, between September and December 2020. The presence of anti IFN-α_2_ Abs was investigated: we first sought to determine a positive cut-off value for Abs detection by performing measurements in 76 putative control sera, *i*.*e*. from healthy donors retrieved before the COVID-19 outbreak. The mean value +3 standard deviation of these measurements provided a cut-off value at 34 ng.mL^-1^. We then assessed the presence of IFN-α2 Abs in putative positive sera, *i*.*e*. sera from patients with autoimmune polyendocrinopathy type 1 syndrome (APS-1), a condition known to be associated with anti-cytokine auto-Abs. All APS-1 patients tested (n=11) had high titers of circulating anti-IFN-α2 auto-Abs (>100ng.mL^-1^). We also evaluated the presence of anti-IFN-α_2_ auto-Abs in 10 mildly-symptomatic COVID-19 healthcare workers and none of them was found positive.

We then measured anti-IFN-α_2_ Abs levels in the sera from the critically-ill COVID-19 patients: 21/84 (25%) were positive and had values above the cut-off (>34 ng.mL^-1^). The neutralizing capacity of their sera against IFN-α was then evaluated as previously described^5^. A neutralizing activity was observed in 15/21 positive sera; in other words, 15/84 (18%) critically-ill COVID-19 patients had neutralizing anti-IFN-α auto-Abs (Figure 1**a**). Importantly, all sera with a titer of anti-IFN-α_2_ auto-Abs above 1 µg.mL^-1^ potently neutralized IFN-α *in vitro*. In addition, in most patients with neutralizing IFN-I Abs, we noticed an impairment of the IFN-I response, which was determined by the measurement of i) plasma IFN-α_2_ levels using the new digital ELISA technology single-molecule arrays (Simoa) and ii) blood Interferon Stimulating Genes (ISG) expression using the Nanostring nCounter technology in blood samples collected in the first 15 days after symptom onset (Figure 1**b-c**). Moreover, there was no significant difference in the clinical characteristics (age, sex ratio, co-morbidity) or outcome (death, O_2_ support) of critically-il COVID-19 patients with or without neutralizing anti-IFN-I auto-Abs (Table 1). Then, serial measurement of IFN-I auto-Abs level during ICU stay was performed for 7 positive patients. The level of IFN-α_2_ auto-Abs remained relatively stable across measurements performed up to 40 days apart (Figure 1**d**). Finally, we assessed the presence of auto-Abs against other IFNs-I in all sera positive for IFN-α2 auto-Abs (n=21). Consistently with previous results^1^, we observed that sera with anti-IFN-α_2_ auto-Ab titers above 1 µg.mL^-1^ also contained auto-Abs against other subtypes of IFN-α and 10/12 contained anti-IFN-ω auto-Abs (Figure 2). Of note, these sera were able to neutralize IFN-ω *in vitro*. Only one serum contained auto-Abs targeting IFN-β at a low titer, and none contained auto-Abs against IFN-ε or IFN-κ.

**Table 1.**
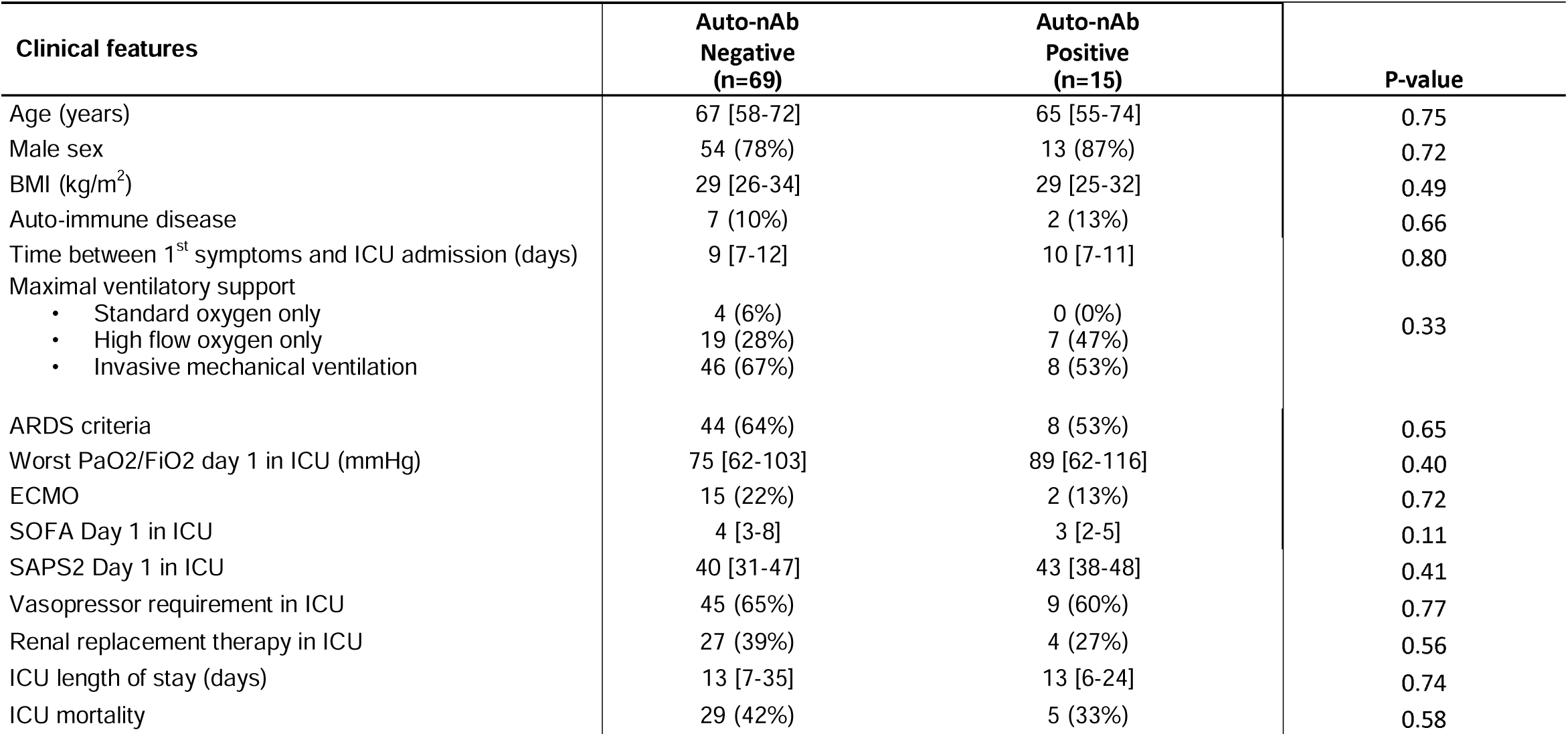
Clinical characteristics of critically-ill COVID-19 patients admitted in intensive care unit. Data are expressed as median [IQR] or count (percentage). Mann-Whitney and Fisher tests were used for quantitative and qualitative variables, respectively. ICU - Intensive Care Unit, BMI - Body Mass Index, nAb – neutralizing Auto-Abs against IFN-α, ECMO-Extracorporeal membrane oxygenation, SOFA - sequential organ failure assessment score, SAPS2 - Simplified Acute Physiology Score II, ARDS - Acute respiratory distress syndrome

| Clinical features | Auto-nAb<br>Negative<br>(n=69) | Auto-nAb<br>Positive<br>(n=15) | P-value |
| --- | --- | --- | --- |
| Age (years) | 67 [58-72] | 65 [55-74] | 0.75 |
| Male sex | 54 (78%) | 13 (87%) | 0.72 |
| BMI (kg/m<sup>2</sup>) | 29 [26-34] | 29 [25-32] | 0.49 |
| Auto-immune disease | 7 (10%) | 2 (13%) | 0.66 |
| Time between 1<sup>st</sup> symptoms and ICU admission (days) | 9 [7-12] | 10 [7-11] | 0.80 |
| Maximal ventilatory support |  |  |  |
| • Standard oxygen only | 4 (6%) | 0 (0%) | 0.33 |
| • High flow oxygen only | 19 (28%) | 7 (47%) |  |
| • Invasive mechanical ventilation | 46 (67%) | 8 (53%) |  |
| ARDS criteria | 44 (64%) | 8 (53%) | 0.65 |
| Worst PaO<sub>2</sub>/FiO<sub>2</sub> day 1 in ICU (mmHg) | 75 [62-103] | 89 [62-116] | 0.40 |
| ECMO | 15 (22%) | 2 (13%) | 0.72 |
| SOFA Day 1 in ICU | 4 [3-8] | 3 [2-5] | 0.11 |
| SAPS2 Day 1 in ICU | 40 [31-47] | 43 [38-48] | 0.41 |
| Vasopressor requirement in ICU | 45 (65%) | 9 (60%) | 0.77 |
| Renal replacement therapy in ICU | 27 (39%) | 4 (27%) | 0.56 |
| ICU length of stay (days) | 13 [7-35] | 13 [6-24] | 0.74 |
| ICU mortality | 29 (42%) | 5 (33%) | 0.58 |

**Figure 1.**
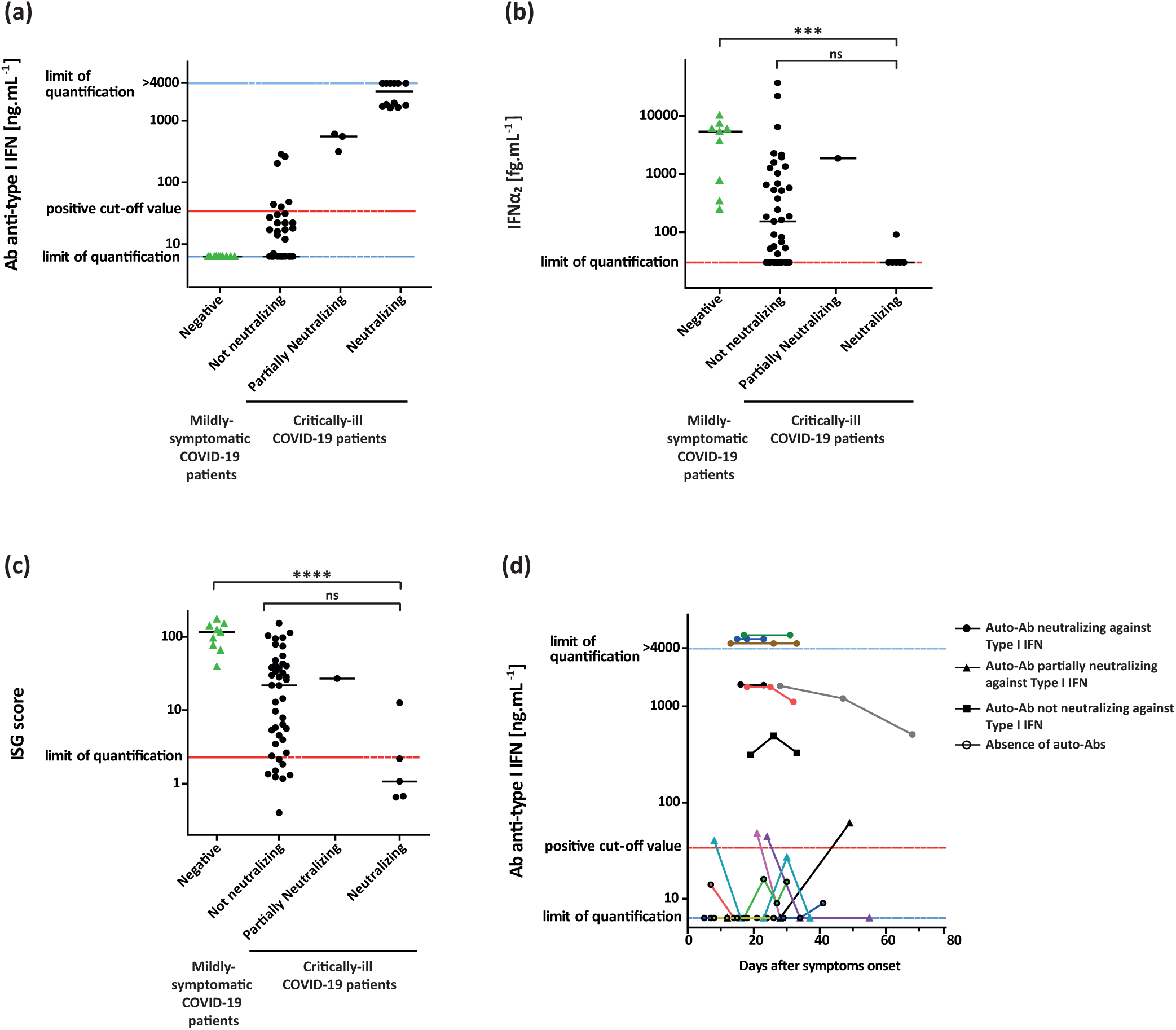
Anti-Type I IFN antibodies (Abs) in patients with life-threatening COVID-19. (a) Concentration of auto-Abs against IFN-α_2_ (ng.mL^-1^) determined by a Thermofisher kit (Catalog # BMS217) in serum samples collected from COVID-19 patients admitted in ICU (n=84) and COVID-19 patients with mild respiratory symptoms (n=10). Auto-Abs concentrations with neutralizing capacity against IFN-α. (b-c) IFN-α_2_ concentration (fg.mL^-1^) (b) and ISG score (c) in plasma and whole blood collected from COVID-19 patients in the first 15 days after symptom onset (critically-ill COVID-19 patients (n=54) and mildly-symptomatic COVID-19 patients (n=10)). (d) Longitudinal detection of auto-Abs against IFN-α_2_ in COVID-19 patients’ serum during their ICU stay according to the delay post-symptom. Dotted lines represent positive cut-off value (threshold), lower limit of quantification (LLOQ), and upper limit of quantification (ULOQ). Solid black lines represent median. Comparisons were performed using the Kruskal Wallis test followed by Dunn’s test. \*\*\**P-*value ≤ 0.001; **** *P-* value ≤ 0.0001.

**Figure 2.**
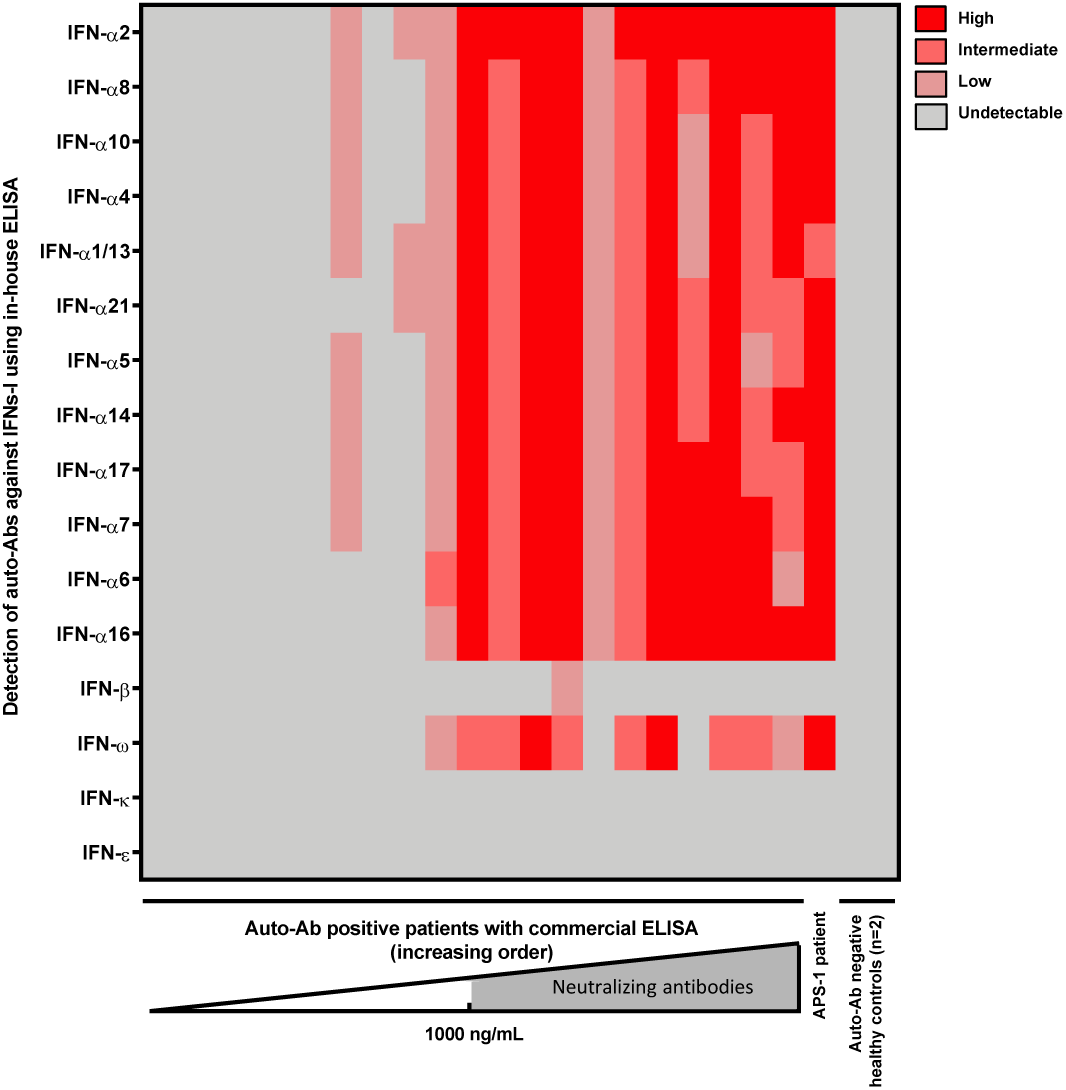
Auto-Abs against other subtypes of IFN-I. The presence of auto-Abs against other subtypes of IFN-I was assessed by ELISA in all sera with anti-IFN-α_2_ auto-Abs (n=21, from left to right, increasing order of concentration of anti-IFN-α_2_). APS-1 patient’s serum was used as positive control and sera from 2 healthy controls were used as negative controls.

## Discussion

A previous study has reported that IFN-I auto-Abs were present in 10.2% of life-threatening COVID-19 patients, undetectable in 663 individuals with asymptomatic or mild COVID-19, and detected in only 0.33% of healthy individuals ^1^. Here, 18% of critically-ill COVID-19 patients were positive for IFN-I auto-Abs whereas all mild-COVID19 patients were negative. We noticed that only a part of auto-Abs detected were able to neutralize IFN-I in the conditions we used, which confirms previous studies in COVID-19 patients and systemic lupus erythematosus subjects ^1, 6^. This finding further confirms the deleterious role of IFN-I autoAbs in the antiviral immune response and the importance of the IFN-I pathway in the defense against SARS-CoV2 infection. Based on its antiviral properties, recombinant IFN-I has been tested as therapy for severe COVID-19, but the treatment showed little or no benefit 7-8. Yet, the potential of such treatment may have been hindered by the presence of IFN-I auto-Abs in patient sera, and this question could therefore be revisited by determining the level of these Abs, for example using the ELISA method we used here. Moreover, patients could be treated with recombinant

IFN-I that are not targeted by auto-Abs (*e*.*g*. IFN-β).

Finally, the detection of anti IFN-I auto-Abs in COVID-19 patients could be useful in routine to identify patients at risk of developing a severe form of the disease. The technique we described here is adequate for this purpose as it can rapidly provide quantitative measurements, and has a cut-off correlated with neutralization assays (1 µg.mL^-1^). However, the presence of auto-Abs was not associated with poorer outcome in critically-ill patients and does not explain all the severe forms of COVID-19, other causes should therefore be sought (*e*.*g*. cytokine release syndrome, presence of other risk factors such as obesity, hypertension).

## Methods

### Participants Critically-ill COVID-19 patients

Plasma samples and Paxgene^®^ tubes were collected from COVID-19 patients hospitalized in the university hospital of Lyon (Hospices Civils de Lyon), France. Diagnosis of COVID-19 was established in all patients by RT-PCR.

All critically-ill patients positive for SARS-CoV-2 virus, admitted to ICU (Croix-Rousse Hospital, Hospices Civils de Lyon), were included in the MIR-COVID study. This study was registered to the *Commission nationale de l’informatique et des libertés* (CNIL, French data protection agency) under the number 20-097 and was approved by an ethics committee for biomedical research (*Comité de Protection des Personnes HCL*) under the number N°20-41. In agreement with the General Data Protection Regulation (Regulation (EU) 2016/679 and Directive 95/46/EC) and the French data protection law (Law n°78-17 on 06/01/1978 and Décret n°2019-536 on 29/05/2019), we obtained consent from each patient or his/her next of kin.

### Mildly-symptomatic COVID-19 patients

Plasma samples and Paxgene^®^ tubes were collected from symptomatic healthcare workers upon COVID-19 diagnosis. Written informed consent was obtained from all participants. The study was approved by the national review board for biomedical research in April 2020 (Comité de Protection des Personnes Sud Méditerranée I, Marseille, France; ID RCB 2020-A00932-37). The study was registered on ClinicalTrials.gov (NCT04341142) where the eligibility, inclusion, and exclusion criteria are previously described ^9^.

### Healthy controls

Prepandemic serum were selected from healthy controls who were recruited among donors to the Lyon blood transfusion centre (Etablissement Français du Sang, EFS). According to French procedures, a written non-opposition to the use of donated blood for research purposes was obtained from HCs. The donors’ personal data were anonymized before transfer to our research laboratory. We obtained approval from the local ethical committee and the French ministry of research (DC-2008-64) for handling and conservation of these samples.

### Auto-Abs anti IFN-I

The presence of anti IFN-α2 auto-Abs was assessed in the plasma using a commercially available kit (Thermo-Fisher, Catalog # BMS217). The positive cut-off value for Ab detection was 34 ng.mL^-1^. The presence of auto-Abs against other IFNs-I was assessed using an ELISA technique, as previously described ^1^. Briefly, 96-well ELISA plates were coated with different cytokines (rhIFN-α2 (Milteny Biotec, ref. number 130-108-984), rhIFN-ω (Merck, ref. number SRP3061) or cytokines from PBL Assay Science (catalog #11002-1), or IFN-β (Milteny Biotech, ref. number: 130-107-888)) and incubated overnight at 4°C. Plates were then washed (PBS 0.005% Tween), then incubated with 5% nonfat milk powder in the same buffer, washed again, and again incubated with 1:50 dilutions of plasma from patients or controls for 2 h at room temperature (or with specific mAbs as positive controls). Horseradish peroxidase (HRP)–conjugated Fc-specific IgG fractions from polyclonal goat antiserum against human IgG, IgM, or IgA (Nordic Immunological Laboratories) were added to a final concentration of 2 μg/mL after thorough washing. Plates were then incubated for 1 h at room temperature and washed. HRP substrate was added and the optical density (OD) was measured.

The neutralization capacity of antibodies against IFN-α2 and –ω was determined as previously described ^5^. Briefly, HEK-293T cells were transfected with a plasmid encoding the firefly Luciferase under the control of human *ISRE* promoters. Cells were then cultured in Dulbecco’s modified Eagle medium (DMEM, Thermo Fisher Scientific) supplemented with 10% healthy control or patient serum/plasma, and were either left unstimulated or were stimulated with IFN-α2, IFN-ω, or IFN-β (10 ng.mL^-1^) for 16 hours at 37°C. Finally, Luciferase level was measured using the Dual-Glo reagent, according to the manufacturer’s instructions (Promega).

### Plasma protein quantification

The concentration of plasmatic IFN-α (fg.mL^-1^) was measured using single molecule array (Simoa) using a commercial kit for IFN-α_2_ quantification (Quanterix(tm), Lexington, MA, USA). The assay was based on a 3-step protocol and an HD-1 Analyzer (Quanterix).

### IFN score assessment

Total RNA was extracted from whole blood stored in Paxgene^®^ tubes (Kit PreAnalytix, Qiagen©, SW) and was quantified using a spectrophotometer (Nanodrop 2000, Thermo Scientific(tm), MA, USA). RNA integrity was then assessed using the Agilent RNA microarray (Agilent Technologies©, Santa Clara, CA, USA). The expression of 6 ISGs (*interferon alpha inducible protein 27* (*IFI27*), *interferon induced protein 44 like* (*IFI44L*), *Interferon Induced Protein With Tetratricopeptide Repeats 1 (IFIT1), ISG15 Ubiquitin Like Modifier (ISG15), Radical S-Adenosyl Methionine Domain Containing 2 (RSAD2), Sialic Acid Binding Ig Like Lectin 1 (SIGLEC1*)) and 3 housekeeping genes (*Actin Beta* (*ACTB), Hypoxanthine Phosphoribosyltransferase 1 (HPRT1), RNA Polymerase II Subunit A (POLR2A*)) was quantified at the transcript level using the Nanostring technology (Nanostring Technologies©, WA, USA). Data standardization was performed using the geometric mean of internal control and housekeeping genes counts. The ISG score was calculated as previously described ^10^.

## Data Availability

The data that support the findings of this study are available from the corresponding author, STA, upon reasonable request.

